# Can we enhance neurorehabilitation through regional implementation of group-based telerehabilitation? A mixed methods evaluation of NeuroRehabilitation OnLine (NROL)

**DOI:** 10.1101/2025.03.10.25323664

**Authors:** Suzanne Ackerley, Thomas Mason, Adam Partington, Rosemary Peel, Helen Vernon, Louise A Connell

## Abstract

**Objectives:** To determine whether neurorehabilitation can be enhanced through regional implementation of group-based telerehabilitation, we implemented the NeuroRehabilitation OnLine (NROL) innovation regionally and evaluated scale-up from a systems perspective.

**Design:** Observational, exploratory evaluation using a mixed methods convergent parallel design.

**Setting:** Stroke and neurological rehabilitation services from four organisations across a regional healthcare system in the United Kingdom.

**Participants:** Therapy staff from community-based services and patients with a stroke or neurological condition receiving active community rehabilitation including NROL from April 2022 to March 2024.

**Intervention:** A regional multidisciplinary group-based neurological telerehabilitation innovation, Neuro Rehabilitation OnLine (NROL).

**Primary outcome measures:** Selected Proctor’s implementation outcomes, to establish system-level adoption, acceptability and sustainability of the regional NROL innovation.

**Results:** NROL was adopted by all intended organisations and continues as part of usual care with participation growing. It was acceptable to therapy staff and patients across the region, well-utilised, valued, and supported increased therapy provision. For sustainability, staffing and travel efficiencies were identified through effective collaborative regional systems working. The importance of continued wide stakeholder engagement, robust evaluation and alignment was highlighted.

**Conclusions:** NROL was successfully embedded into real-world practice at a system-level and enhanced neurorehabilitation. Looking forward, longer-term sustainment of this innovation will require a compelling business case and value proposition for decision-makers, addressing economic, equality, and operational efficiency considerations.

**ARTICLE SUMMARY:** *Strengths and limitations of this study:*

- Within a clinical-academic partnership, robust mixed methods evaluation enabled comprehensive system-level insights into the scale-up of a group-based telerehabilitation innovation
- Our approach was strengthened by using established implementation science outcomes and frameworks
- Use of real-world routinely collected data was pragmatic and integral to the model’s embedded nature but limited the focus of insight predominantly to those who participated in NROL
- Decision-maker perspective was not formally captured but will be an important next step, alongside gaining opinion from those who did not participate

## INTRODUCTION

Multidisciplinary rehabilitation plays an important role in minimising disability, maximising function, and optimising quality of life for people living with long-term neurological conditions. Higher doses of neurorehabilitation improve outcomes (1–4). In England, despite agreed recommendations for therapy amount (5, 6), actual provision falls significantly below recommended levels, with variable access across providers (7); a pattern seen internationally (8, 9). Barriers to guideline adherence include workforce challenges, inefficiencies in resource allocation and lack of supporting organisational processes (10–12). Achievement of recommended targets for access and provision may remain unmet until transformative system-level change is undertaken (11, 12), however solutions need to be feasible within our existing healthcare settings.

Telerehabilitation, the delivery of rehabilitation remotely via telecommunication devices, is one solution that may help address shortfalls in therapy provision and is an approach endorsed for use alongside conventional in-person therapy (5). When delivered remotely, rehabilitation interventions have been shown to be safe and equally effective as those delivered in-person including for stroke and neurological populations for patient outcomes such as activities of daily living and health-related quality of life (13–16). A scoping review indicated that limited technical skills hinders telerehabilitation implementation whilst predominant facilitators include patient motivation and leadership involvement (17). Patient and clinician uptake and satisfaction is facilitated by appropriate training (technology use, supporting clinicians’ adaptation of practice for remote delivery) and the use of telerehabilitation to augment rather than replace in-person therapy (18). Telerehabilitation can minimise barriers to access, such as reducing travel requirement and optimising resource use, and supports standardisation of care whilst facilitating rehabilitation engagement (19–21). These advantages may be especially important in areas with widely dispersed populations, a lack of local services, and/or staffing shortages. As national healthcare services face reform, strategic drivers aim to shift resources to the community, improve productivity and flow, and integrate digital transformation initiatives (22, 23). Incorporating acceptable telerehabilitation models into existing care pathways and systems may provide opportunity for enhancing rehabilitation with efficient resource use (24).

NeuroRehabilitation OnLine (NROL) is a multidisciplinary, group-based, real-time telerehabilitation innovation that was successfully adapted from a pilot version (25) and implemented at a single National Health Service (NHS) community stroke and neurological service in the United Kingdom (UK) (19). It leverages group-based therapy, which is supported by favourable evidence, benefits peer-support and offers staffing efficiencies (19, 25). Embedded within the service’s established neurorehabilitation care pathway, NROL contributed to the rehabilitation offer, sitting alongside existing therapy provision as part of a hybrid model of care. Supported by a dedicated small operational team, including technology support, it offered advantages to save time, energy and travel, enabling remote therapy to be delivered using existing workforce (19). NROL at this single-service level was deemed appropriate for patients with various neurological conditions and acceptable to patients and therapy staff, with positive outcomes and opinions. The innovation aligned with strategic priorities, including the use of data and digital technologies in healthcare (26). NROL was acknowledged in UK and Ireland stroke guidelines as an exemplar innovation for remotely delivered rehabilitation (5). To build on these positive results, a decision was made to expand NROL into a regional innovation (27, 28).

Scale-up of NROL was a ‘deliberate effort to increase the impact of this successful innovation to a greater number of services across the region’ (29), and aligned with statutory changes for healthcare organisations to work cross-regionally to improve population healthcare, tackle unequal outcomes and access, and enhance productivity and value for money (30). Scaling up efforts should be underpinned by key principles including systems thinking and a focus on sustainability (29, 31).

Within a clinical-academic partnership, we sought to evaluate whether our regional community neurorehabilitation services could be enhanced through system-level implementation of the group-based telerehabilitation programme, NROL. Our objective was to evaluate the scale-up of NROL, embedded within established stroke and neurological rehabilitation pathways across multiple services at four healthcare organisations, from a system-level perspective. Implementation success was conceptualised by exploring the following questions:

As a regional innovation:

1) Was NROL adopted across the region?
2) Was NROL accepted and a way to support increased therapy provision?
3) What key considerations help inform NROL’s sustainability?

## METHODS

Real-world implementation of NROL was examined through an observational, exploratory, service evaluation. A mixed methods design was used and is described below, preceded by an overview of context. This approach is particularly valuable when evaluating the implementation of complex innovations, where both measurable outcomes and contextual factors matter (32). Manuscript preparation was guided by the Good Reporting of a Mixed Methods Study (GRAMMS)(33).

Implementation was explored using selected Proctor’s implementation outcomes of adoption, acceptability and sustainability (34), agreed with decision-makers (NHS managers and commissioners) as relevant, and within resources available. System-level indicators for the implementation outcomes were:

- Adoption: NROL participation and delivery across organisations, and challenges and enablers to adoption.
- Acceptability: NROL utilisation, safety and perceived value for therapy staff and patients, and a way to support increased therapy provision.
- Sustainability: Key considerations for NROL to continue as usual care as a regional innovation, including model efficiencies (staffing, travel avoidance) and sustained appropriateness (representativeness of NROL participants to the patient population, NROL patient outcomes consistent with single-service NROL evaluation).

### Implementation context

The regional NROL innovation was co-produced within a collaboration between a regional healthcare system (Lancashire and South Cumbria Integrated Care System) and a clinical-academic partnership (Lancaster University and East Lancashire Hospitals NHS Trust (ELHT)). ELHT, the host healthcare organisation, registered the NROL project as a service evaluation, with data routinely collected and anonymised. Implementation was guided by the updated Medical Research Council (MRC) framework for developing complex interventions (28, 32). From April 2022, a horizontal phased approach (29) scaled NROL from a single-service (ELHT) to a multi-service model, integrating three further organisations (University Hospitals of Morecambe Bay NHS Foundation Trust (UHMB), Blackpool Teaching Hospitals NHS Foundation Trust (BTH) and Lancashire and South Cumbria NHS Foundation Trust (LSCFT)) (28). Context was described using the Consolidated Framework for Implementation Research (CFIR), a commonly used determinant framework providing a menu of constructs under domain headings that have been associated with effective implementation (35, 36). A summary is provided below under relevant domain headings, with full details published separately (27, 28).

### Inner setting (Healthcare setting)

Community-based stroke and neurological rehabilitation services from four organisations (ELHT, UHMB, BTH, LSCFT) within a regional healthcare system in the UK were involved. These services provide multidisciplinary rehabilitation for adults with sudden onset or progressive/intermittent neurological conditions. Before NROL’s scale-up (April 2022), services existing rehabilitation offer primarily consisted of in-person individual evidence-based therapy at patients’ home, with limited onsite group and occasional online individual therapy sessions. The exception was ELHT, who had provided online group therapy via NROL at a single-service level since January 2021.

### Innovation (NROL)

NROL as a regional innovation has been comprehensively described (27). Embedded into established neurorehabilitation care pathways, the NROL programme aimed to improve access to therapy by providing telerehabilitation to complement the existing rehabilitation offer. In brief, NROL delivered real-time (synchronous) group-based multidisciplinary evidence-based therapy online (Microsoft Teams) over repeating six-week ‘NROL blocks.’ Each NROL block began with an entry session, participants then followed individualised programmes tailored to their needs, including weekly targeted talking (e.g. cognitive, communication, well-being) and/or physical (e.g. mobility, upper limb) therapy groups and an optional community peer-support group, for six-weeks. Groups were delivered by therapy staff (allied health professionals, psychologists, assistants), patient volunteers and students. To facilitate participation, patients received set-up and ongoing assistance from a dedicated NROL Technology Support team member, who was present during sessions. Clinical reasoning determined if and when an NROL programme fitted within a patient’s overall rehabilitation. Patients typically exited the programme at the end of their 6-week block, however could attend more than one NROL block if clinically indicated providing they remained under the active care of their stroke or neurorehabilitation team.

### Individuals (Patients and staff)

Patients undertaking community neurorehabilitation could be referred to NROL by therapy staff. In addition to service eligibility, inclusion required English proficiency (or translator access) and willingness to participate in online group therapy, with or without carer support. Participants required access to suitable computer hardware or connectivity, though support was sought from Age UK. Patients remained part of a local treating therapist’s caseload who was ultimately responsible for their overall rehabilitation. Existing therapy staff from all involved services delivered NROL collectively (i.e. shared care approach), coordinated by a dedicated regional NROL team (Operational manager, Technology Support, Administrator). Clinical-academics and organisational (NHS) neurorehabilitation managers provided leadership.

### Outer setting (External influences)

NROL aligned with UK national healthcare strategic drivers including an ambition for equitable regional service delivery, managing workforce challenges, efficient resource use, digital innovation and delivering towards environmental targets (22, 23, 26, 37–39), and is endorsed in UK and Ireland stroke guidelines (5).

### Design

A mixed-methods convergent parallel design was used to evaluate NROL scale-up and is overviewed in Figure 1. Service data (quantitative) were collected throughout the evaluation period (April 2022-March 2024), with NROL service data analysed concurrently. Survey data (quantitative and qualitative) were collected from service staff (June – July 2023) and NROL patients (April 2022 – August 2023) and analysed separately. Results relating to the system-level indicators were synthesised and interpreted under selected implementation outcomes.

**Figure 1.**
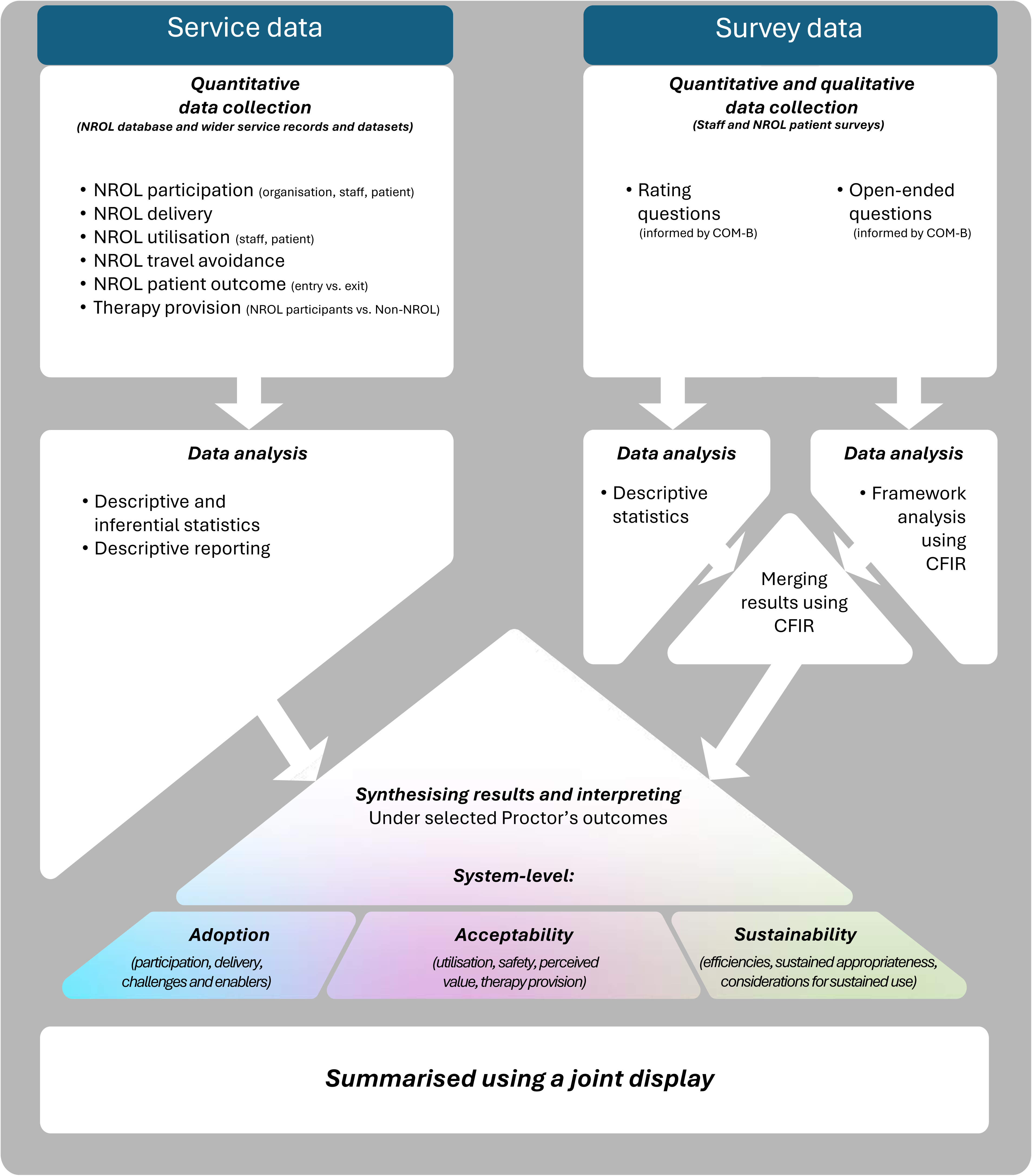
NROL evaluation mixed methods convergent parallel design. Service and survey data were collected and analysed separately. Results relating to the system-level indicators were synthesised under selected implementation outcomes and summarised using a joint display. CFIR = Consolidated Framework for Implementation Research (36). COM-B = Capability, Opportunity, Motivation-Behaviour Framework (40).

### Data collection and analysis

Table 1 provides an overview of the evaluation components to demonstrate data alignment with relevant implementation outcomes and system-level indicators, and to outline the analysis approach. Additional details are provided below and in Supplementary file 1.

**Table 1:**
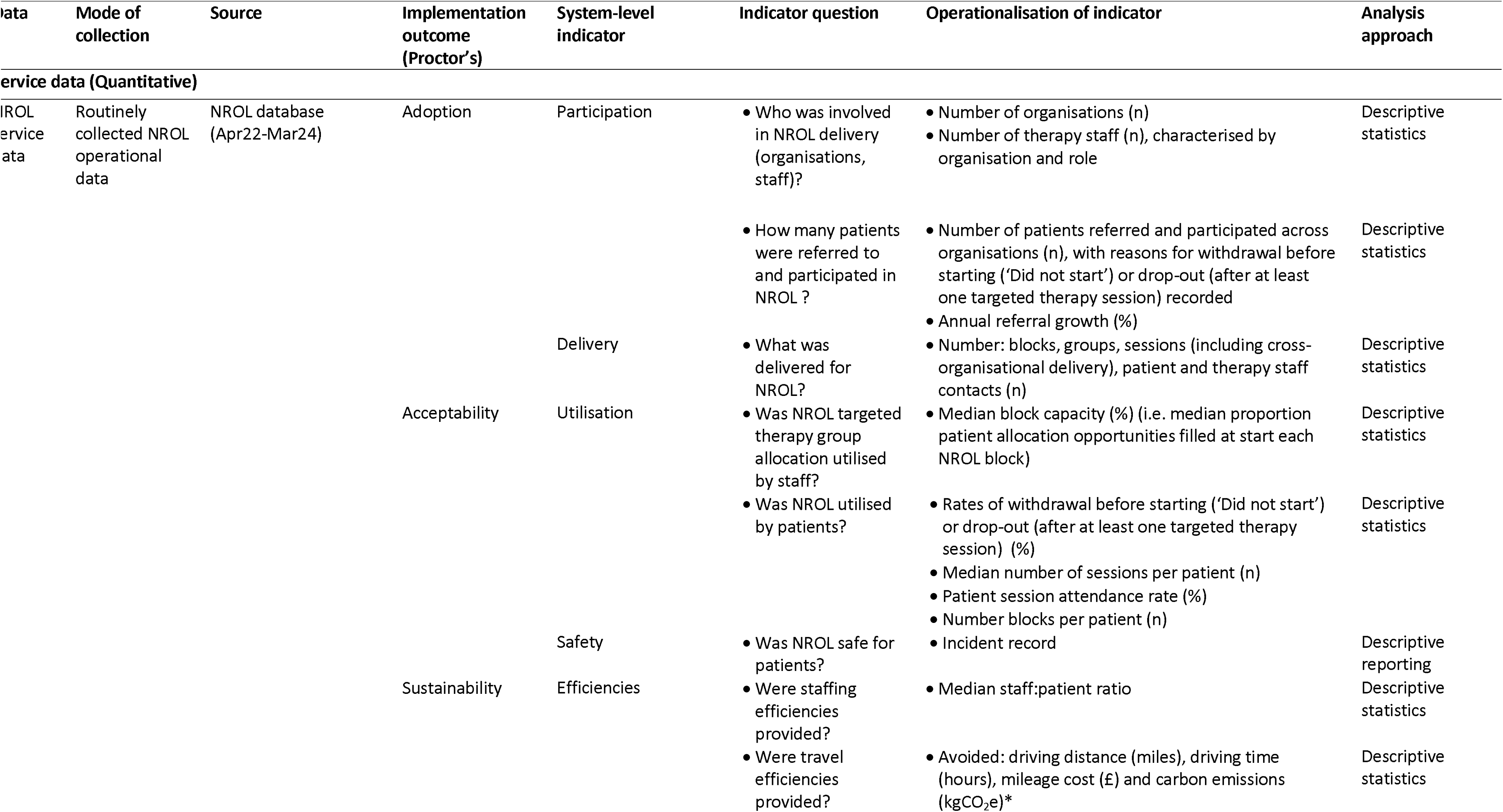
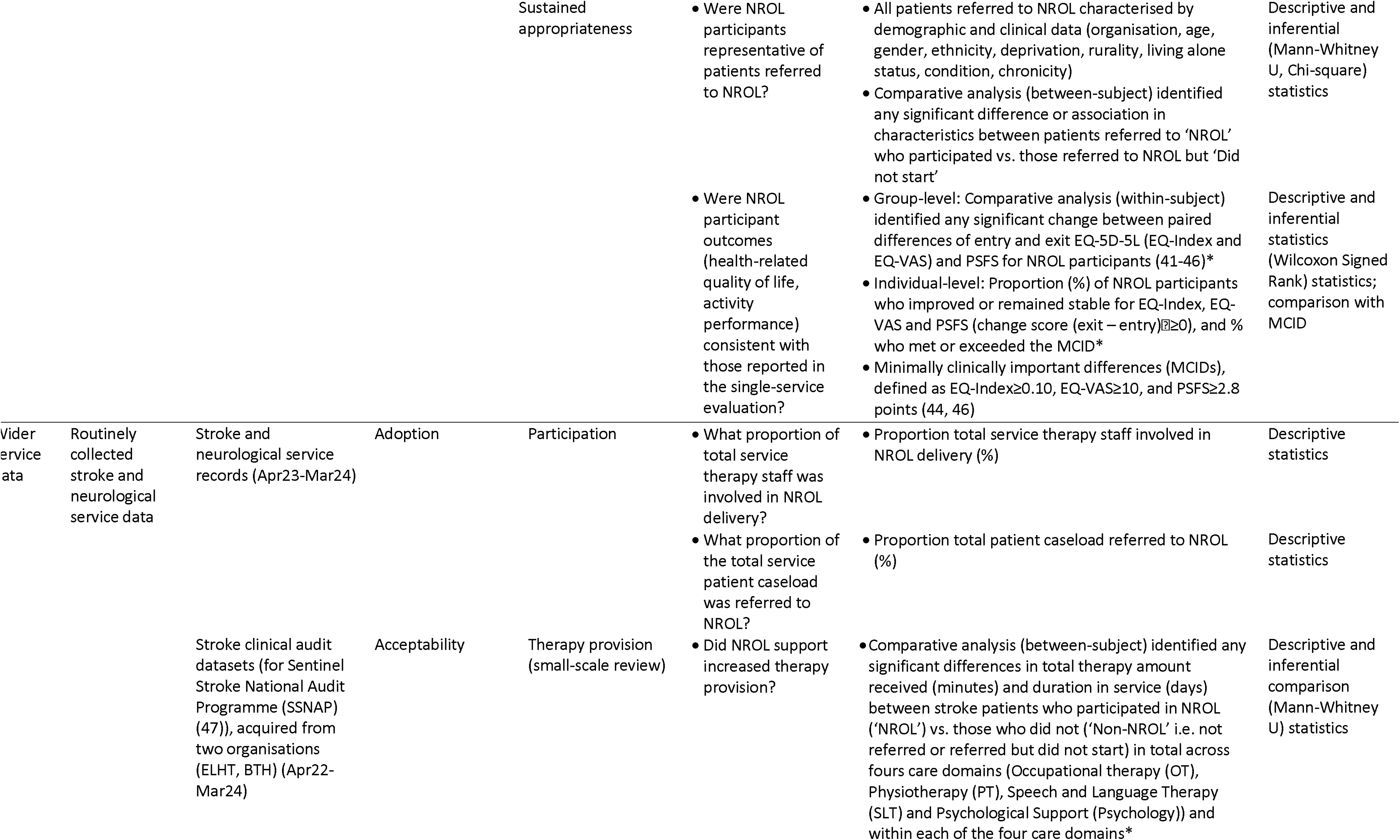
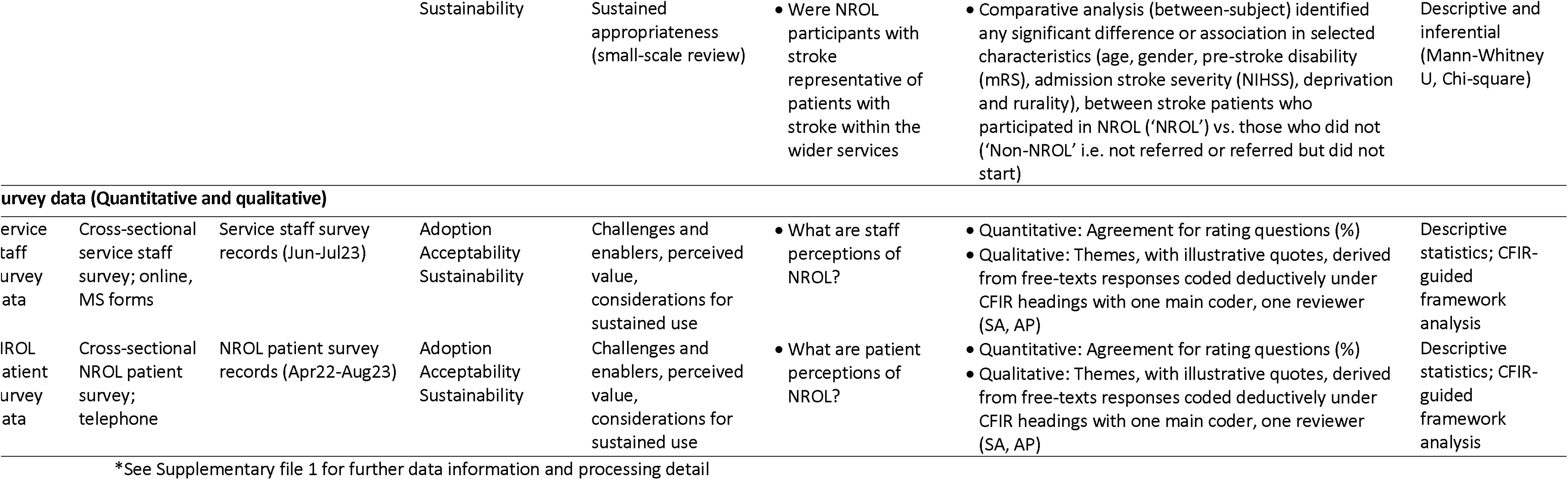
Overview of evaluation components.

Service data were obtained from 1) NROL service data from the NROL database, inputted by the NROL operational team and group facilitators and 2) relevant wider service data from service records and stroke clinical audit datasets, provided retrospectively by service managers where possible. For NROL service data, intention to treat analysis was applied for participants who dropped-out (i.e. after at least one targeted therapy session). Service data were predominantly summarised using descriptive statistics and reporting. Comparative analysis with inferential statistical testing was used to quantify therapy provision between patients with stroke who participated in NROL (‘NROL’) and those who did not (‘Non-NROL’ i.e. wider service patients not referred to NROL or referred but did not start NROL). It was also used to evaluate sustained appropriateness (representativeness of NROL participants to the patient population, NROL patient outcomes (within-subject) consistent with single-service NROL evaluation (19)). See Table 1 for detail.

Survey data were obtained from cross-sectional surveys undertaken by service staff and NROL patients within participating services across the region as part of service evaluation. Survey development was informed by the COM-B (Capability, Opportunity, Motivation-Behaviour) framework (40) and related to project outcomes. The COM-B was chosen as it recognises key factors integral in changing behaviour, is widely used in public health, and aligns with the updated CFIR used for analysis. The surveys used a mix of multiple choice, Likert-scale rating, and open-ended questions. The open-ended questions were used to provide deeper insights into implementation challenges and enablers, perceptions of NROL’s value, and key considerations for its sustained use. They were each piloted on 2-3 people to check content, clarity, usability and time taken and minimally refined to address feedback (see Supplementary file 2 for final versions). The staff survey link was shared to all relevant service staff via service management to collect anonymous feedback. Patients were surveyed via telephone as part of their NROL exit procedure and were aware that survey administrator was not a member of the clinical team, and an honest perspective was wanted. Survey data were analysed using descriptive statistics for quantitative rating responses and CFIR-guided framework analysis for qualitative free-text responses. For each stakeholder group, quantitative and qualitative survey findings were merged and summarised under relevant CFIR-headings to detail system-level constructs influencing implementation, with equal emphasis placed on both strands.

Analyses were performed using Microsoft Excel (Version 2411), StataNow/MP 18.5, and IBM SPSS Statistics version 29.0. All statistical tests were two-tailed, and statistical significance was determined at a p-value threshold of 0.05. Where normality testing (Shapiro-Wilk test) indicated significant non-normality (p<0.05), nonparametric statistical testing was used and medians, with ranges or interquartile ranges (IQR), reported.

### Mixed methods synthesis

A joint display (48) was used to provide structure to summarise the service and survey analyses under the selected implementation outcomes, a recommended approach for presenting mixed methods results (49) and consistent with our prior reporting (19). Aligning service results with survey findings, enabled synergistic insights when exploring system-level adoption, acceptability and sustainability. Insights were summarised by SA and LC and discussed and agreed by the clinical authors (AP, RP, HV) and the wider stakeholders involved in NROL.

### Evaluator characteristics and reflexivity

This project was part of a clinical academic partnership, with LC & SA being both experienced researchers and physiotherapists in neurorehabilitation who have been involved in NROL development, implementation, evaluation, and sustainment. AP is a physiotherapist who works as the operational manager of NROL for Lancashire and South Cumbria. RP is an occupational therapist at UHMB who delivers NROL. HV is a speech and language therapist and Head of Service for Stroke and Neurorehabilitation at ELHT and provides strategic leadership for NROL. TM is an applied econometrician working as a quantitative researcher focused on health and health care. The evaluators’ embedded role in NROL implementation provided valuable contextual insight. However, they remained mindful of potential bias and took steps to mitigate this. Evaluation was guided by logic models and informed by regular engagement with diverse stakeholders. Findings and interpretations were discussed and agreed with the wider stakeholder groups involved in NROL.

### Patient and public involvement (PPI)

Stakeholder engagement is a core element in developing and evaluating complex interventions (32). Patients and carers co-designed NROL within a learning collaborative, providing vital contributions to enhance its quality and relevance as previously described with a GRIPP2 reporting checklist (27). Involvement has continued throughout implementation, with patients providing feedback to refine content and delivery. Engagement has resulted in tangible improvements, for example a trial of a ‘Vocation’ group and the creation of patient video vignettes to communicate key NROL information to diverse audiences in an accessible and meaningful way.

## RESULTS

Service data is reported over the total evaluation period (April 2022–March 2024). Supplementary file 1 contains tables presenting data annually, and from the preceding year of single-service NROL (April 2021–March 2022, ELHT), which enabled review of NROL delivery, utilisation and efficiencies over time.

NROL surveys were completed by 49 therapy staff (estimated response rate 36% of regional community neurorehabilitation therapy workforce) and 176 NROL patient (response rate 85% of NROL participants completing NROL in the survey time window). Respondents’ characteristics (Supplementary file 2:Table) reflect regional therapy workforce and NROL patient characteristics and participation profile (Supplementary file 1:Tables1.1,1.7, Figure 2).

**Figure 2:**
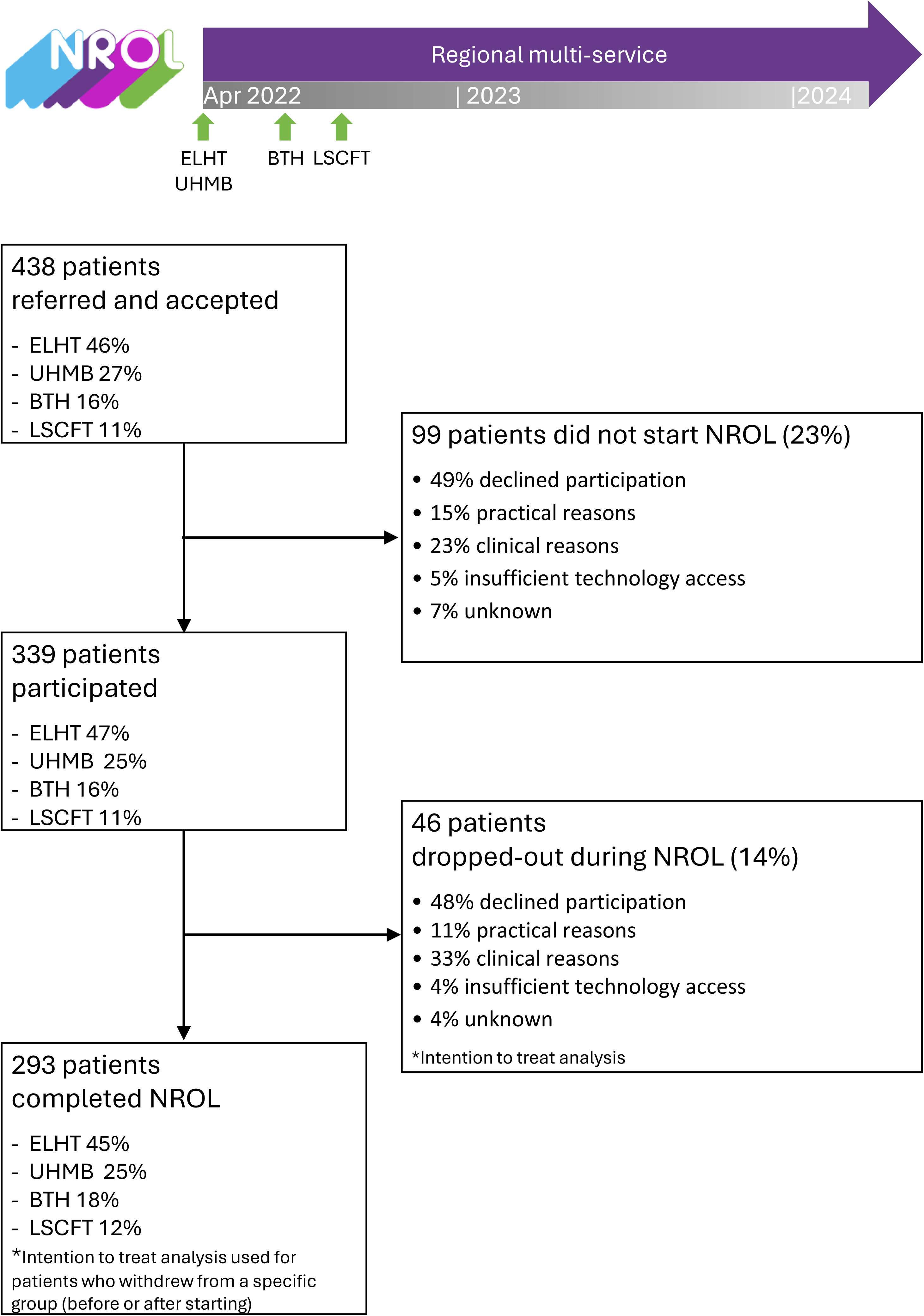
Regional NROL organisation and patient participation profile. Profiles of participation across community-based stroke and neurological rehabilitation services of the four organisations within the regional healthcare system (East Lancashire Hospitals NHS Trust (ELHT), University Hospitals of Morecambe Bay NHS Foundation Trust (UHMB), Blackpool Teaching Hospitals NHS Foundation Trust (BTH) and Lancashire and South Cumbria NHS Foundation Trust (LSCFT)).

Results are presented according to the system-level indicators for each implementation outcome.

### System-level adoption: participation, delivery, challenges and enablers

#### NROL participation

Phased NROL adoption by all four organisations was completed across a 6-month period between April – September 2022 (Figure 2). During the evaluation period, 75 existing multidisciplinary therapy staff from across the four organisations delivered NROL groups within their existing role (Supplementary file 1:Table1.1 for role and organisation detail), supported by three NROL operational team members. Per NROL block, about 40 therapy staff (∼30% regional community neurorehabilitation therapy workforce) delivered sessions. Additionally, 167 staff registered to observe NROL. The NROL patient participation profile is depicted in Figure 2: 438 patients were referred, reflecting approximately 2– 16% of each organisation’s wider service patient caseload (Supplementary file 1:Table1.2), with annual referral growth (Single-Regional Y1: 59%; Regional Y1-Y2: 30%, Supplementary file 1:Table1.3); 339 patients participated (77%), with 293 (67%) of referred patients completing their programme.

#### NROL delivery

Key delivery data are summarised in Table 2, with further data (including group-level data) provided in Supplementary file 1 (Supplementary Tables:1.3,1.4). Twelve NROL blocks were run successfully involving ten different groups (Cognitive education, Cognitive processing, Adjustment/well-being, Fatigue, Dysarthria, Aphasia, Vocation, Balance and Mobility, Upper limb, Peer-support). Sixty-nine percent of the sessions were run cross-organisationally (i.e. therapy staff from ≥2 organisations). The proportion of therapy staff contacts from each organisation ranged from 7% to 53% (variation likely reflecting phased organisational participation and different service sizes), however when considered relative to referral proportion differences were between-6% and 7%. To support delivery, there were 488 NROL technology support contacts, 23 student, and 19 patient-volunteer (for peer-support group). NROL remains part of usual care.

**Table 2:** NROL delivery, utilisation and efficiency summary.

|  | Apr 22–Mar 24 |
| --- | --- |
| Service metric | Regional Total |
| **Total: Delivery and staffing efficiency** |  |
| Blocks delivered, n | 12 |
| Sessions delivered, n | 511 |
| Patient contacts, n | 3434 |
| Therapy staff contacts, n | 1167 |
| Therapy staff:patient ratio, median (range) | 0.4 (0.1 – 2.0) |
| **Targeted therapy groups: Utilisation** |  |
| Block capacity, median % | 84% |
| Patient session attendance rate, % | 71% |

#### Challenges and enablers to adoption

Survey findings are summarised in Table 3, detailing system-level key constructs identified as influencing NROL implementation. Content includes challenges and enablers to adoption, along with perceived value and future considerations as presented in subsequent result subsections.

**Table 3:**
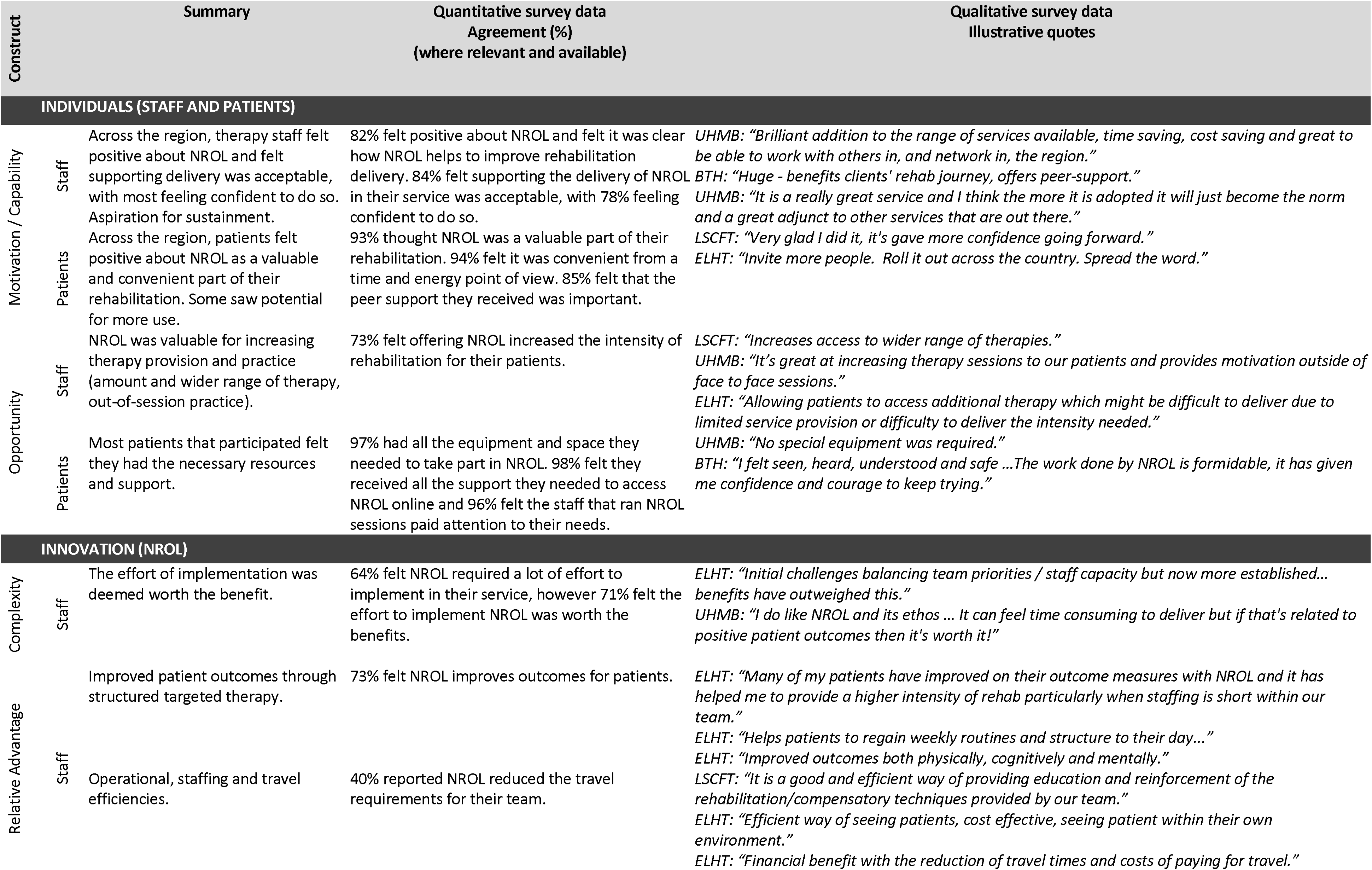
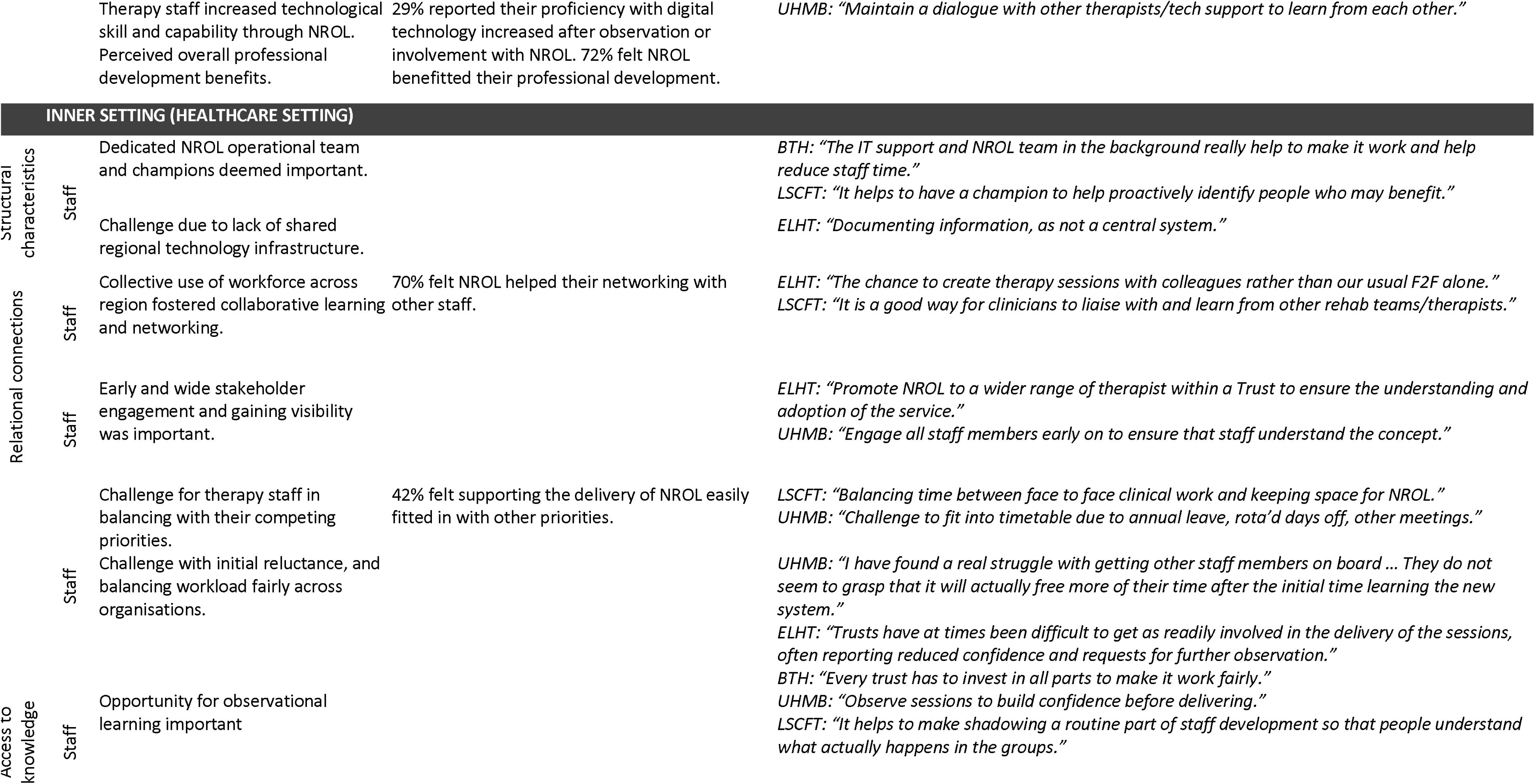
System-level key constructs influencing NROL implementation.

Challenges to adoption included limited shared technological infrastructure, initial reluctance and difficulty balancing workload alongside competing priorities and across organisations. Staff also recognised that implementation took up-front effort, but that demands lessened as familiarity increased. Key enablers identified included staff confidence aided by observational opportunity to access knowledge about NROL, and structural characteristics such as leadership from a dedicated operational team (including technology support) and innovation champions. These ‘Inner setting’ constructs were reported by staff only, as patients are typically less aware of healthcare system factors.

### System-level acceptability: utilisation, safety, perception of NROL, and supported increased therapy provision

#### NROL utilisation

Key utilisation data for the targeted therapy groups are summarised in Table 2, with further detail (including group-level data) provided in Supplementary file 1 (Supplementary Tables:1.3,1.4). Blocks were run at a median of 84% capacity. NROL withdrawal (did not start) and drop-out (after at least one targeted therapy session) rates were 23% and 14% respectively (Figure 2), and across organisations ranged from 20–28% and 2-18%. Participants attended a median of 9 sessions (range = 1–50). Overall targeted therapy patient session attendance rate was 71% (group range 60–83%), with organisation patient session attendance rate ranging between 65–83%. The peer-support group was optional to attend, and 200 patients (59%) chose to participate. Most patients participated in one NROL block (79%), but some participated in two (18%), three (2%) or four (1%).

#### Safety

One non-injury event was recorded: a minor seizure relating to a known condition during a talking session. NROL staff followed standard operating procedure, no first aid was required and the patient was able to continue participation.

#### Perceptions

Survey findings (Table 3) demonstrated concordant positive feedback. The majority of therapy staff felt positive about NROL and found supporting delivery acceptable. NROL was perceived by many as a valuable opportunity for increasing therapy provision, practice and outcomes, with some identifying efficiencies. Implementation benefits were considered worthwhile to justify the effort to adopt. Additional value was perceived through professional development (including technology skills), as well as collaborative learning and networking. Patients across the region indicated high satisfaction with NROL. It was seen as a valuable and convenient part of their rehabilitation and they felt they had the necessary resources and support providing opportunity to take part. Both staff and patients reported motivational benefits, including the value of peer support.

#### Therapy provision

Figure 3 illustrates the distribution of total therapy provision for patients with stroke that were ‘NROL’ participants and ‘Non-NROL’ patients. Small-scale review (n: NROL= 76, Non-NROL= 1,520) demonstrated patients with stroke who participated in NROL received significantly more therapy (Median therapy minutes received (IQR): NROL=1535 (1015–2855) vs. Non-NROL=335 (165–833), p<0.001). This reflected a significant increase in receipt of therapy for OT and PT domains (both p<0.001). Additionally, patients with stroke who participated in NROL had a longer duration in service (Median days (IQR): NROL=146 (117–168) vs. Non-NROL=59 (28–109), p<0.001); with increased duration for the allied health professions (OT, PT, SLT all p<0.05)). Full details across the four care domains are provided in Supplementary file 1 (Supplementary Table:1.5).

**Figure 3:**
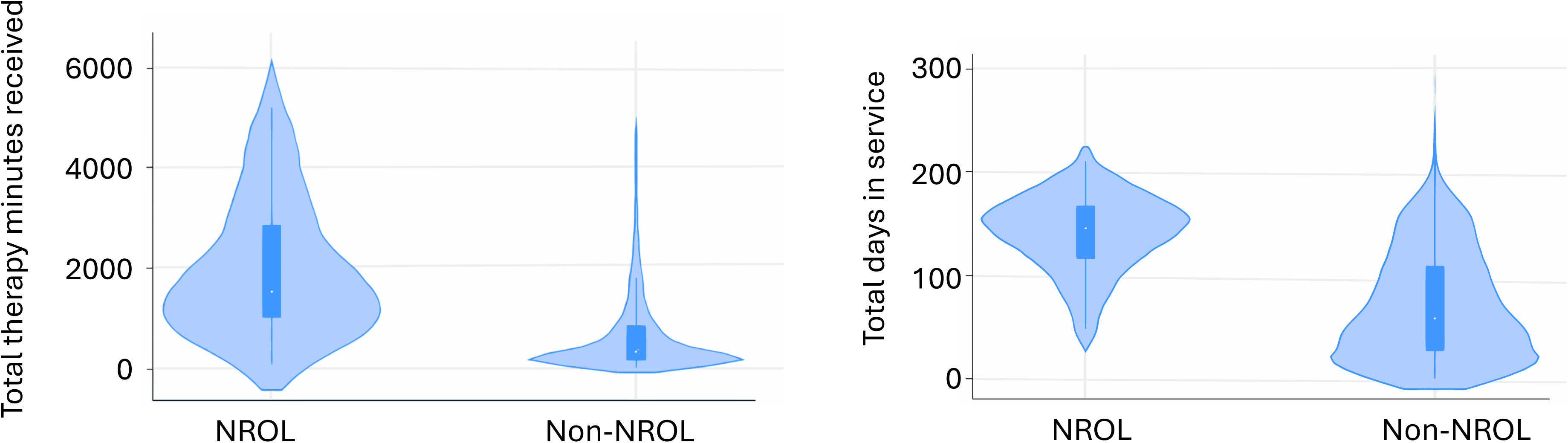
Violin plots of therapy provision. Violin plots showing distributions of total therapy minute received (A) and total days in service (B). Representations are for participants with stroke who received NROL as part of their rehabilitation (‘NROL’) and those who did not (‘Non-NROL’). Plots show the range, interquartile range, median, and the density (or frequency) of data points across values, with wider sections indicating more participants with data at those values.

### System-level sustainability: efficiencies, sustained appropriateness, and future considerations

#### Efficiencies

Staffing efficiencies were achieved, with a median staff:patient ratio of 0.4 across all sessions (Table 2), improving annually from 0.4 to 0.3 (Supplementary file 1:Table1.3). Group-level data demonstrated that all groups were delivered with a median ratio less than or equal to 0.8 (Supplementary file 1:Table1.4).

For travel avoidance, avoided driving miles totalled 75554 (Annual average=37777 miles) and avoided driving time 2506 hours (annual average=1253 hours). Avoided mileage cost totalled £44577 (annual average=£22289). Avoided carbon emissions totalled 23046 kgCO_2_e (annual average=11523 kgCO_2_e). For annual details see Supplementary file 1 (Supplementary Table:1.6).

#### Sustained appropriateness

Representativeness of NROL participants to the patient population

Participating patient characteristics are presented in Table 4, demonstrating varied patients accessed NROL. Patients’ residences were across an area circa 4480km^2^ , with the percent of regional NROL participants living rurally (23%) aligning with that of the region (20%) (50). Compared to patients that were referred to NROL but did not start, patients that participated in NROL were younger (Median age, y (IQR): NROL=60 (50–71) than those who were referred but did not start=63 (55–74), p<0.05) and more likely to have conditions that were sudden onset (p<0.01) and at the subacute phase (p<0.05). Small-scale review of selected characteristics indicated patients with stroke who participated in NROL were younger than those who did not participate (Median age (IQR): NROL=61 (54–71) vs. Non-NROL=75 (64–82), p<0.01). They also statistically had lower level pre-stroke disability (Mean mRS: NROL=0.41 (1.01) vs. Non-NROL=0.87 (1.19), p<0.01), however both were less than 1 (where 0 indicates no symptoms and 1 indicates no significant disability and normal functioning) and the difference may be consistent with age differential. See Supplementary file 1 (Supplementary Tables:1.7,1.8) for all comparisons.

**Table 4:**
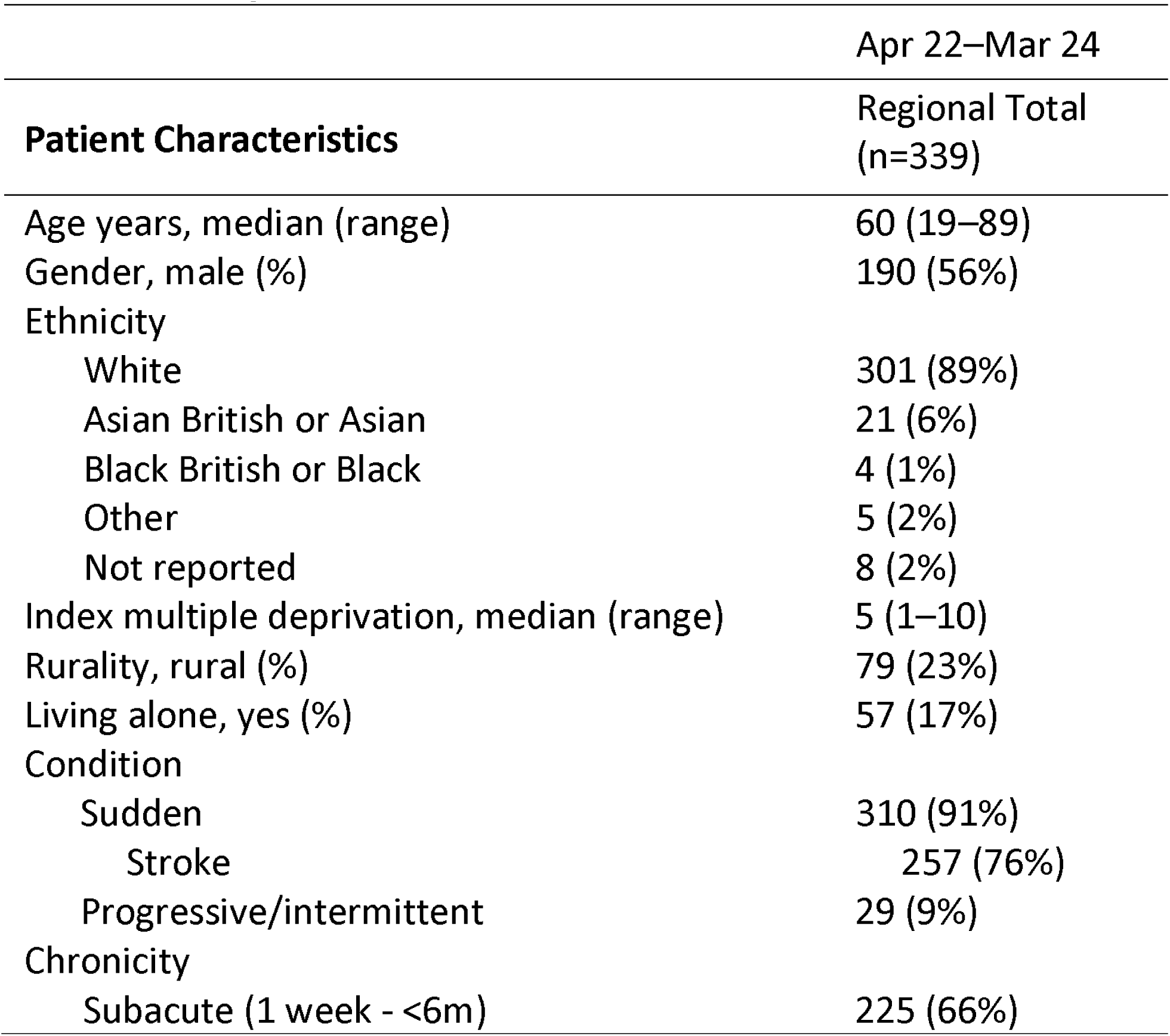
NROL patient characteristics.

|  | Apr 22–Mar 24 |
| --- | --- |
| **Patient Characteristics** | **Regional Total**<br>**(n=339)** |
| Age years, median (range) | 60 (19–89) |
| Gender, male (%) | 190 (56%) |
| Ethnicity |  |
| White | 301 (89%) |
| Asian British or Asian | 21 (6%) |
| Black British or Black | 4 (1%) |
| Other | 5 (2%) |
| Not reported | 8 (2%) |
| Index multiple deprivation, median (range) | 5 (1–10) |
| Rurality, rural (%) | 79 (23%) |
| Living alone, yes (%) | 57 (17%) |
| Condition |  |
| Sudden | 310 (91%) |
| Stroke | 257 (76%) |
| Progressive/intermittent | 29 (9%) |
| Chronicity |  |
| Subacute (1 week - <6m) | 225 (66%) |

#### NROL patient outcomes

Complete data sets were available for EQ-5D-5L and PSFS for 280 and 282 patients (83%) respectively. At a group level, statistically significant improvements were observed over time for both health-related quality of life measures (EQ-Index, EQ-VAS) and activity performance (PSFS)(all p<0.01). Large overlaps in interquartile ranges and standard deviations were noted and no difference met the minimally clinically important difference (MCID). Overall, these results indicate a systematic positive shift in paired differences, rather than a substantial change in central tendency. At an individual level, the proportions of NROL participants who improved or remained stable were between 64%-80%, with 30%-39% of participants meeting or exceeding the MCID. For full details see Supplementary file 1 (Supplementary Table:1.9).

#### Future considerations

Survey findings (Table 3) highlights staff and patient aspiration for NROL sustainment and further use. Staff indicated the importance of relational connections, with early and wide stakeholder engagement and gaining visibility deemed important.

### Mixed methods synthesis

A joint display (Figure 4) shows the convergence across service and survey analyses to explore the implementation outcomes of system-level adoption, acceptability and sustainability. Insights from the wider NROL stakeholders gathered through collaborative discussions shaping this paper highlighted the value of capturing and sharing robust evaluation and feedback, and underscored the necessity for strategic alignment across multiple levels. These perspectives have been incorporated into sustainability content.

**Figure 4.**
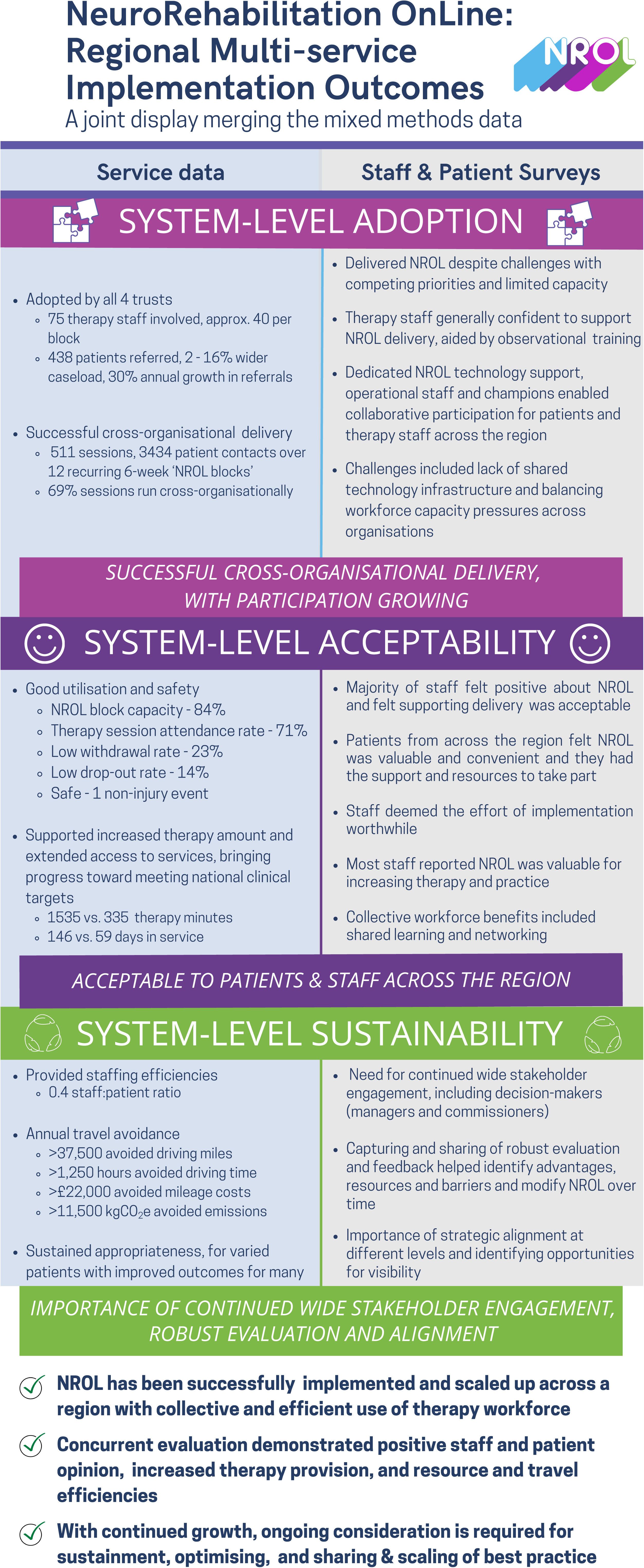
Joint display merging mixed methods data.

## DISCUSSION

This observational mixed methods evaluation of the system-level scale-up of NROL, a group-based telerehabilitation innovation, demonstrated enhancement of our regional community neurorehabilitation services. Despite anticipated healthcare system pressures, NROL was widely adopted and successfully embedded within real-world practice at all intended organisations across the region, with growing participation supported by collaborative system-level working. Acceptability was evidenced by the programme being well-utilised and valued by patients and therapy staff, while safely supporting increased therapy provision. Successful adoption, acceptability, and model efficiencies indicate strong potential for sustained delivery, although continued stakeholder engagement and alignment remain essential for long-term sustainability.

A data-driven approach enabled real-time tailoring to optimise innovation-context fit, an important factor in scaling efforts (29, 51). NROL proved compatible at a system-level, with phased adoption across three additional organisations over six months. This led to growth, with increased referrals in the first year (59%) followed by annual growth (30%), whilst demonstrating improved staffing efficiency over time (staff:patient ratio 0.4 to 0.3). Despite this growth, referrals accounted for a relatively small proportion of the wider caseload (2–16%), indicating potential for further participation. However, with block capacity nearing maximum (84%) broader participation may require model adjustments.

Implementation was influenced by individual and healthcare setting factors, such as initial therapy staff reluctance, competing priorities and infrastructure challenges, consistent with reported barriers to scale-up, innovation implementation and sustained use in healthcare (52, 53). Nevertheless, key enablers facilitated success, in particular leadership from a dedicated operational team and innovation champions. The importance of highly committed individuals is well-recognised (52–56). The impetus to overcome barriers may have been influenced by staffs’ recognition of the potential long-term benefits for patients, including improved outcomes for many and peer-support. Consistent with staff perceptions and previous single-service NROL evaluation (19), quantitative results indicated positive shifts in activity performance and health-related quality of life, supporting the programme’s sustained appropriateness when delivered at a regional level. In line with the prior findings, clinically meaningful group-level differences were not detected, although meaningful gains were reported for some individuals (30-39%). This may not be surprising given the relatively short duration of the NROL programme (typically six-weeks). From October 2024 the EQ-5D-5L has been included within the national community stroke clinical audit (e.g. SSNAP, 47) which may enable longer-term, and comparative, patient outcome review within future evaluation. Notably, staff valued NROL for increasing therapy and practice. Quantitative data supported their views, showing patients with stroke who participated in NROL received over than four times as many therapy minutes as non-NROL patients and were supported by community services for longer, marking significant steps toward meeting therapy targets.

Workforce limitations remain a major barrier to implementation and sustainability in healthcare systems (52, 57). We employed collective use of the regional rehabilitation therapy workforce, as a shared care approach, to help mitigate this challenge. This approach yielded several advantages. Firstly, it allowed a critical mass of patients to receive group therapy where impairment incidence was lower (e.g. speech-related groups). The cross-organisational structure (69% of sessions run cross-organisationally) reduced the need for multiple therapy staff from any one service per session, while providing opportunity for shared learning and networking. It also offered redundancy, allowing staff shortages in one service to be temporarily covered by another, minimising regional inequity. Although a concern regarding workload balance across organisations was voiced, staffing distribution aligned relatively well with referral patterns (variation –6% to 7%) suggesting a natural equilibrium or effective management. The collaborative system-level approach was enabled by the online format, which had the additional advantages of travel time, cost and emissions avoidance. Further, overall therapy staff resource use was minimised through the group-based model. Together, these advantages may help alleviate pressure on the healthcare system.

Wide stakeholder engagement fostered buy-in and short-term sustainment of the regional NROL innovation. Therapy staff and patients were integral to successful development and implementation as shown here and seen previously (19, 27). However, regular communication with decision-makers was also crucial for scale-up, though this perspective is yet to be formally captured as it was beyond the resource of this evaluation. Alignment (i.e. creating fit between elements of the inner and outer setting of an organisation or system) plays an important role for implementation success (58, 59). Anecdotally, decision-makers found NROL appealing due to its strategic alignment with community-based care priorities, its low operational resource use, and its contribution to broader healthcare goals, including workforce planning, performance benchmarks (e.g. SSNAP), environmental targets and addressing health inequalities. However, sustained funding is essential to prevent “pilotitis” and ensure long-term viability.

Some limitations should be considered when interpreting the findings. First, the therapy provision review was small-scale and limited to patients with a diagnosis of stroke, as clinical audit data were unavailable for other neurological conditions. Also, we only acquired access to stroke clinical audit datasets from two organisations. Further, interpretation should consider that the patients with stroke who participated in NROL may have been those who needed, could manage, or were motivated for therapy, potentially affecting therapy amount and duration. Second, the staff survey had a 36% estimated response rate (55% respondents had delivered NROL), introducing potential nonresponse bias. This is not unexpected, as subject familiarity increases survey participation likelihood. However, it is important to note that those with less positive experience may be underrepresented. However, a strength of a mixed-methods approach is that the use of qualitative and quantitative data can help illustrate converging or diverging factors, providing a more comprehensive view. Finally, it was difficult to capture what was not done (e.g. who was not referred). Whilst overall NROL had sustained appropriateness for mixed demographics, we found patients referred to NROL and went on to participate were younger. Factors could be gate-keeping from therapists or patient access to technology; this requires further exploration. Going forward these perspectives will be sought, as well as the aforementioned perspective of decision-makers.

## CONCLUSION

NROL was adopted and embedded into usual care at a regional level as an adjunct to enhance neurorehabilitation. It was accepted at a system-level and increased the amount of therapy provided to patients. Workforce efficiencies were identified through effective collaborative systems working, with benefits amplified as participation grew. Longer-term sustainment of this regional innovation will require a compelling business case and value proposition for decision-makers. Ongoing work will address broader economic factors, focus on equality, and improve operational efficiencies while supporting environmental goals. Importantly, adjustments to the model will be explored to optimise capacity and further intensify the therapy offer.

## Data Availability

The datasets used and/or analysed during this evaluation are available from the corresponding author on reasonable request.

## ABBREVIATIONS

BTH: Blackpool Teaching Hospitals NHS Foundation Trust
CFIR: Consolidated Framework for Implementation Research
ELHT: East Lancashire Hospitals NHS Trust
EQ-5D-5L: EuroQol 5-Dimensions 5-Level
GRAMMS: Good Reporting for A Mixed Methods Study
GRIPP2: Guidance for Reporting Involvement of Patients and the Public
LCSFT: Lancashire and South Cumbria NHS Foundation Trust
MCID: Minimal Clinically Important Difference
MRC: Medical Research Council
mRS: Modified Rankin Scale
NHS: National Health Service
NIHSS: National Institutes of Health Stroke Scale
NROL: NeuroRehabilitation OnLine
OT: Occupational Therapy
PSFS: Patient Specific Functional Scale
PT: Physiotherapy
SLT: Speech and Language Therapy
UHMB: University Hospitals of Morecambe Bay NHS Foundation Trust
UK: United Kingdom

## ACKNOWLEDGEMENTS

We thank the following who contributed to the regional NROL innovation: Staff, patients and carers from the stroke and neurological rehabilitation services at East Lancashire Hospitals NHS Trust, University Hospitals of Morecambe Bay NHS Foundation Trust, Blackpool Teaching Hospitals NHS Foundation Trust and Lancashire and South Cumbria NHS Foundation Trust. Ian Grimshaw and Katie McGoogan for providing technological and administrative support to staff and patients. We convey our appreciation to SameYou charity for their ongoing partnership to support the NROL programme and their generous fundraising.

## AUTHOR CONTRIBUTIONS

SA and LC made substantial contributions to the conception and design of the work; SA, AP, TM and LC made substantial contributions to the acquisition, analysis and interpretation of the data; SA and LC drafted the work. All authors provided critical input and revised the manuscript draft. All authors have read and approved the final version.

