## Supplementary tables 1 for "Can we enhance neurorehabilitation through regional implementation of group-based telerehabilitation? A mixed methods evaluation of NeuroRehabilitation OnLine (NROL)"

**Supplementary file 1: Service data tables**

Includes Supplementary:

- Table 1.1: NROL therapy staff and regional community neurorehabilitation therapy workforce characteristics
- Table 1.2: Referrals to NROL as a proportion of referrals to the wider service patient caseload by organisation
- Table 1.3. NROL delivery and utilisation summary over time
- Table 1.4: NROL group delivery and attendance over 12 blocks regional NROL
- Table 1.5: Therapy provision for stroke patients: NROL participants vs. Non-NROL patients
- Table 1.6: NROL Travel avoidance
- Table 1.7. NROL patient characteristics over time
- Table 1.8: Selected characteristics of stroke patients: NROL participants vs. Non-NROL patients
- Table 1.9: NROL patient outcomes

**Supplementary Table 1.1: NROL therapy staff and regional community neurorehabilitation therapy workforce characteristics**

| | **Evaluation period**<br>**Regional**<br>**Apr 22–Mar 24[a]** | **Apr 23-Mar 24[b]** |
|---|---|---|
| **Characteristics** | **NROL**<br>**Regional therapy staff (%)**<br>**(N=75)** | **Wider community neurorehabilitation services**<br>**Regional therapy workforce (%)**<br>**(N=135)** |
| **Organisation** | | |
| ELHT | 41% | 40% |
| UHMB | 29% | 16% |
| BTH | 19% | 23% |
| LSCFT | 11% | 21% |
| **Role** | | |
| Occupational therapist | 35% | 27% |
| Physiotherapist | 33% | 27% |
| Psychology | 4% | 1% |
| Speech and Language Therapist | 12% | 15% |
| Dietician | 1% | 1% |
| Therapy assistants/trainees | 15% | 28% |

Regional = multi-service. [a]2-year evaluation period (Regional Total). [b]Annual period

**Supplementary Table 1.2: Referrals to NROL as a proportion of referrals to the wider service patient caseload by organisation**

| | Evaluation period<br>Regional<br>Apr 22–Mar 24[a] |
|---|---|
| | % NROL referrals relative to wider service patient caseload |
| **Organisation** | |
| ELHT | 7% |
| UHMB | 16% |
| BTH | 2% |
| LSCFT | 2% |

**Supplementary Table 1.3: NROL delivery and utilisation summary over time**

| | Annual Single-service Apr 21–Apr 22 | Annual Regional Apr 22–Mar 23 | Apr 23–Mar 24 | **Evaluation period Regional Apr 22–Mar 24** |
|---|---|---|---|---|
| **Service metric** | Single-service (S)[a] | Regional 1 (R1)[a] | Regional 2 (R2)[a] | **Regional Total[b]** |
| **Total: Targeted therapy and community groups, entry session** | | | | |
| Blocks delivered, n | 6 | 6 | 6 | 12 |
| Sessions delivered, n | 198 | 255 | 256 | 511 |
| Patients referred, n | 121 | 192 | 249 | 438 (ELHT 46%, UHMB 27%, BTH 16%, LSCFT 11%) |
| Patients participated, n | 98 | 155 | 187 | 339 (ELHT 47%, UHMB 25%, BTH 16%, LSCFT 11%) |
| Patient contacts, n | 1057 | 1618 | 1816 | 3434 (ELHT 45%, UHMB 28%, BTH 16%, LSCFT 11%) |
| Median patients participating per block, n (range) | 21 (17-26) | 32 (27 -36) | 38 (30 – 50) | 34 (27-50) |
| Median sessions per participating patient, n (range) | 8 (1-35) | 10 (1-47) | 8 (1-50) | 9 (1–50) |
| Patient session attendance rate, % | 58% | 58% | 55% | 56% |
| Therapy staff contacts, n | 361 | 585 | 582 | 1167 (ELHT 53%, UHMB 29%, BTH 10%, LSCFT 7%) |
| Therapy staff:patient ratio, median (range) | 0.4 (0.1–3.0) | 0.4 (0.1–2.0) | 0.3 (0.1–2.0) | 0.4 (0.1–2.0) |
| **Targeted therapy groups: Talking and Physical** | | | | |
| Groups delivered per block, median (range) | 5 (4–6) | 7 (6–7) | 7 (5–8) | 7 (5–8) |
| Sessions delivered, n | 163 | 214 | 214 | 428 |
| Patients participated, n | 96 | 153 | 184 | 334 |
| Patient contacts, n | 739 | 1091 | 1252 | 2343 |
| Median patients per group, n (range) | 4 (1-11) | 5 (1-12) | 6 (1-13) | 5 (1-13) |
| Median block capacity[c], % | 73% | 80% | 85% | 84% |
| Patient session attendance rate, % | 67% | 69% | 72% | 71% (ELHT 65%, UHMB 73%, BTH 78%, LSCFT 83%) |
| Therapy staff contacts, n | 293 | 496 | 466 | 962 |
| Therapy staff:patient ratio, median (range) | 0.5 (0.1–3.0) | 0.5 (0.1–2.0) | 0.4 (0.1–2.0) | 0.4 (0.1–2.0) |
| **Community: Peer-support group[d,e]** | | | | |
| Sessions delivered, n | 29 | 35 | 36 | 71 |
| Patients participated, n | 64 (65%) | 97 (63%) | 104 (56%) | 200 (59%) |
| Patient contacts, n | 256 | 417 | 437 | 854 |
| Median patients per group, n (range) | 9 (4-14) | 12.5 (6-21) | 12.5 (6-20) | 12 (6-21) |
| Patient session attendance rate, %[e] | 41% | 38% | 32% | 35% |
| Therapy staff contacts, n | 54 | 72 | 99 | 171 |
| Therapy staff:patient ratio, median (range) | 0.2 (0.1–0.4) | 0.2 (0.1–0.4) | 0.2 (0.1–0.5) | 0.2 (0.1–0.5) |

Regional = multi-service. [a]Annual periods (S, R1, R2). [b]2-year evaluation period (Regional Total). Three patients participated in both annual reporting periods (R1 and R2) and are included in each. [c]% Block capacity reflects proportion of patient allocation opportunities filled at start of NROL block. [d]NROL entry sessions are only reflected in total. [e]Peer-support group is optional attendance for all patients.

**Supplementary Table 1.4: Group delivery and attendance over 12 blocks regional NROL**

| | Evaluation period Regional Apr 22-Mar24[a] | | | | | | | |
|---|---|---|---|---|---|---|---|---|
| **Group/s** | **Sessions delivered, n** | **Patients participated, n** | **Proportion patients in group, %** | **Patient contacts** | **Median group capacity, % (target group size)** | **Patient session attendance rate, %** | **Therapy staff contacts** | **Therapy Staff:patient ratio, median (range)** |
| **Total (incl. Peer-support group)** | 511 | 339 | 100% | 3434 | | 56% | 1167 | 0.4 (0.1 – 2.0) |
| **Total (excl. Peer-support group)** | 440 | 339 | 100% | 2580 | | 71% | 996 | 0.4 (0.1 – 2.0) |
| **Total Targeted therapy** | 428 | 334 | 99% | 2343 | | 71% | 962 | 0.4 (0.1 – 2.0) |
| **Talking** | 287 | 250 | 74% | 1426 | | 71% | 619 | 0.5 (0.1 – 2.0) |
| Cognitive education | 72 | 64 | 19% | 293 | 92% (6) | 74% | 163 | 0.6 (0.3 – 1.5) |
| Cognitive processing | 35 | 38 | 11% | 124 | 83% (6) | 69% | 78 | 0.7 (0.3 – 2.0) |
| Adjustment/wellbeing | 32 | 79 | 23% | 183 | 58% (12) | 71% | 68 | 0.4 (0.2 – 1.0) |
| Fatigue | 83 | 136 | 40% | 608 | 100% (12) | 67% | 173 | 0.3 (0.1 – 0.8) |
| Dysarthria | 39 | 27 | 8% | 119 | 67% (6) | 81% | 85 | 0.8 (0.3 – 2.0) |
| Aphasia | 25 | 23 | 7% | 96 | 83% (6) | 83% | 50 | 0.5 (0.2 – 1.0) |
| Vocation | 1 | 3 | 1% | 3 | 83% (6) | 60% | 2 | 0.7 (0.7 – 0.7) |
| **Physical** | 141 | 138 | 41% | 917 | | 70% | 343 | 0.4 (0.2 – 1.5) |
| Balance & Mobility | 72 | 97 | 29% | 535 | 88% (12) | 72% | 201 | 0.4 (0.2 – 0.8) |
| Upper Limb | 69 | 80 | 24% | 382 | 71% (12) | 67% | 142 | 0.4 (0.2 – 1.5) |
| **Total Community** | 83 | 286 | 84% | 1091 | | 40% | 205 | 0.2 (0.1 – 0.5) |
| NROL entry session | 12 | 236 | 70% | 237 | | 81% | 34 | 0.2 (0.1 – 0.2) |
| Peer-support[b] | 71 | 200 | 59% | 854 | | 35% | 171 | 0.2 (0.1 – 0.5) |

Regional = multi-service. [a]2-year evaluation period (Regional Total). [b]Peer-support group is optional to attend with all patients invited to all sessions.

**Therapy provision**

**Data information and processing:**

- Stroke clinical audit datasets (for Sentinel Stroke National Audit Programme (SSNAP)), acquired from two organisations (ELHT, BTH)
- All available stroke patient records within the evaluation period were included, except patients who didn't access community services.

**Supplementary Table 1.5: Small-scale review therapy provision for stroke patients: NROL participants vs. Non-NROL patients**

| | | **Evaluation period Regional Apr22-Mar24** | | | | | | | | | | |
|---|---|---|---|---|---|---|---|---|---|---|---|---|
| | | **NROL** | | | | | **Non-NROL** | | | | | **p-value[a]** |
| | | n | Mean | Median | SD | IQR | n | Mean | Median | SD | IQR | |
| Total | Total duration of stay in service (days) | 76 | 140.22 | 146.00 | 38.34 | 117-167.5 | 1,520 | 71.02 | 59.00 | 53.24 | 28-109 | **p<0.001** |
| | Total therapy received (minutes) | 76 | 2017.34 | 1535.00 | 1371.94 | 1015-2855 | 1,520 | 694.77 | 335.00 | 902.63 | 165-832.5 | **p<0.001** |
| OT | Duration in service in OT rehabilitation (days) | 68 | 122.51 | 133.50 | 48.04 | 87.5-157.5 | 1,086 | 65.02 | 53.00 | 50.63 | 24-97 | **p<0.001** |
| | Total OT received (minutes) | 68 | 753.74 | 755.50 | 556.46 | 300-995 | 1,086 | 288.34 | 175.00 | 342.92 | 90-335 | **p<0.001** |
| PT | Duration in service in PT rehabilitation (days) | 69 | 111.49 | 121.00 | 57.06 | 65-156 | 1,106 | 69.24 | 61.00 | 52.62 | 22-108 | **p<0.001** |
| | Total PT received (minutes) | 69 | 1097.99 | 845.00 | 996.54 | 320-1625 | 1,106 | 507.15 | 257.50 | 639.45 | 120-615 | **p<0.001** |
| SLT | Duration in service in SLT rehabilitation (days) | 29 | 94.28 | 90.00 | 64.46 | 32-146 | 508 | 63.16 | 48.00 | 54.04 | 19-95 | **0.011** |
| | Total SLT received (minutes) | 29 | 505.97 | 80.00 | 885.22 | 45-525 | 508 | 303.54 | 95.00 | 534.85 | 45-280 | 0.712 |
| PSY | Duration in service in psychological support rehabilitation (days) | 31 | 102.23 | 107.00 | 64.56 | 41-154 | 131 | 94.64 | 82.00 | 63.77 | 42-155 | 0.507 |
| | Total psychological support received (minutes) | 31 | 375.16 | 150.00 | 373.94 | 60-680 | 131 | 212.25 | 120.00 | 227.63 | 60-300 | 0.082 |

Stroke patient data only. Fours care domains = Occupational therapy (OT), Physiotherapy (PT), Speech and Language Therapy (SLT) and Psychological Support (PSY). [a]Mann-Whitney U test. NROL = NROL participants. Non-NROL patients = not referred to NROL or referred but did not start.

**NROL Travel avoidance**

**Data information and processing:**

- Informed estimates of avoided driving miles and time were determined using Google maps, based on referring staff worksite and patient residential postcode combined with NROL session attendance
- Mileage cost avoidance 59 pence per mile (NHS mileage rate, NHS Employers)
- Avoided carbon emissions estimated using total avoided mileage cost and a UK conversion factor (0.517 $kgCO^2e$ per £, DEFRA)

**Supplementary Table 1.6: NROL Travel avoidance**

| | Annual Regional Apr 22–Mar 23 | Apr 23–Mar 24 | **Evaluation period Regional Apr 22–Mar 24** |
|---|---|---|---|
| **Travel metric** | Regional 1 (R1)[a] | Regional 2 (R2)[a] | **Regional Total[b]** |
| Avoided driving miles, n | 38364 | 37191 | 75554 |
| Avoided driving time, hours | 1254 | 1252 | 2506 |
| Avoided mileage cost, £ | 22635 | 21942 | 44577 |
| Avoided carbon emissions, $kgCO_2e$ | 11702 | 11344 | 23046 |

Regional = multi-service. [a]Annual periods (R1, R2). [b]2-year evaluation period (Regional Total).

**Supplementary Table 1.7. NROL patient characteristics over time**

| | Annual Single-service | Annual Regional | | **Evaluation period Regional** | | |
|---|---|---|---|---|---|---|
| | Apr 21–Apr 22 | Apr 22–Mar 23 | Apr 23–Mar 24 | **Apr 22–Mar 24** | | |
| **Characteristics** | Single-service (S)[a] Participants (N=98) | Regional 1 (R1)[a] Participants (N=155) | Regional 2 (R2)[a] Participants (N=187) | **Regional Total[b] Participants (n=339)** | **Regional Total[b] Did not start (n=99)** | **P-value[c]** |
| Organisation | | | | | | |
| ELHT | 98 (100%) | 80 (52%) | 82 (44%) | 161 (47%) | 40 (40%) | |
| UHMB | | 46 (30%) | 41 (22%) | 85 (25%) | 33 (33%) | |
| BTH | | 16 (10%) | 38 (20%) | 55 (16%) | 14 (14%) | |
| LSCFT | | 13 (8%) | 26 (14%) | 38 (11%) | 12 (12%) | |
| Age years, median (range) | 57 (19–86) | 60 (20–89) | 60 (19–89) | 60 (19–89) | 63 (22-89) | **0.023** |
| Gender, male (%) | 58 (59%) | 90 (58%) | 100 (54%) | 190 (56%) | 50 (51%) | 0.302 |
| Ethnicity | | | | | | |
| White | 73 (75%) | 140 (90%) | 164 (88%) | 301 (89%) | 91 (92%) | |
| Asian British or Asian | 10 (10%) | 8 (5%) | 13 (7%) | 21 (6%) | 5 (5%) | |
| Black British or Black | - | 2 (1%) | 2 (1%) | 4 (1%) | - | |
| Other | - | 3 (2%) | 2 (1%) | 5 (2%) | - | |
| Not reported | 15 (15%) | 2 (1%) | 6 (3%) | 8 (2%) | 3 (3%) | |
| Index multiple deprivation, median (range) | 4 (1–10) | 5.5 (1–10) | 5 (1–10) | 5 (1–10) | 5 (1 – 10) | 0.670 |
| Rurality, rural (%) | 18 (18%) | 42 (27%) | 38 (20%) | 79 (23%) | 17 (17%) | 0.196 |
| Living alone, yes (%) | 14 (14%) | 26 (17%) | 31 (17%) | 57 (17%) | 25 (26%) | 0.053 |
| Condition | | | | | | **0.009** |
| Sudden | 73 (75%) | 137 (88%) | 175 (94%) | 310 (91%) | 98 (99%) | |
| Stroke | 45 (46%) | 109 (70%) | 150 (80%) | 257 (76%) | 85 (86%) | |
| Progressive/intermittent | 25 (26%) | 18 (12%) | 12 (6%) | 29 (9%) | 1 (1%) | |
| Chronicity | | | | | | |
| Subacute (1 week - <6m) | 46 (47%) | 89 (57%) | 138 (74%) | 225 (66%) | 79 (80%) | **0.011** |

Single = single-service, Regional = multi-service. [a]Annual periods (S, R1, R2). [b]2-year evaluation period (Regional Total). Three patients participated in both annual reporting periods (R1 and R2) and are included in each. [c]Mann-Whitney U or Chi-square. Participants with missing values for a given variable were excluded from that specific analysis only; missingness was minimal (all <2%).

**Supplementary Table 1.8. Small-scale review of selected characteristics for stroke patients: NROL participants vs. Non-NROL patients**

**Data information and processing:**

- Stroke clinical audit datasets (for Sentinel Stroke National Audit Programme (SSNAP)), acquired from two organisations (ELHT, BTH)
- All available stroke patient records within the evaluation period were included, except patients who didn't access community services.

| | **Evaluation period Regional Apr22-Mar24** | | | | | | | | | | |
|---|---|---|---|---|---|---|---|---|---|---|---|
| | **NROL** | | | | | **Non-NROL** | | | | | |
| **Characteristic** | **n** | **Mean** | **Median** | **Min** | **Max** | **n** | **Mean** | **Median** | **Min** | **Max** | **p-value[b]** |
| Age, years | 76 | 61.03 | 60.50 | 31 | 89 | 1520 | 72.71 | 75.00 | 18 | 100 | **<0.001** |
| Gender, % male[a] | 76 | 57.89 | - | 0 | 1 | 1520 | 56.58 | - | 0 | 1 | 0.821 |
| Pre-stroke disability, mRS | 56 | 0.41 | 0.00 | 0 | 4 | 1267 | 0.87 | 0.00 | 0 | 5 | **<0.001** |
| Admission stroke severity, NIHSS | 56 | 6.80 | 4.00 | 0 | 28 | 1267 | 6.21 | 4.00 | 0 | 42 | 0.386 |
| Index multiple deprivation | 76 | 4.46 | 4.00 | 1 | 10 | 1506 | 4.24 | 4.00 | 1 | 10 | 0.413 |
| Rurality, % rural[a] | 76 | 7.89 | - | 0 | 1 | 1505 | 14.22 | - | 0 | 1 | 0.120 |

Stroke patient data only. [a]Median values not provided for binary variables. [b]Mann-Whitney U or Chi-square. NROL = NROL participants. Non-NROL = patients not referred to NROL or referred but did not start.

**NROL patient outcomes**

**Data information and processing:**

- Health-related quality of life was measured by the EQ-5D-5L consisting of two parts: the EQ descriptive system profiles individual's health state and EQ Visual Analogue Scale (EQ-VAS, max 10) rates individual's perceived overall current health (EQ-5D-5L, Golicki et al. 2015, Chen et al. 2016). Descriptive system question responses were transformed into an EQ Index score (EQ-Index, max 1.000) using the EuroQoL Group's crosswalk methodology with a United Kingdom population value set (van Hout et al. 2012)
- Activity performance measured by the Patient Specific Functional Scale (PSFS, max 10), was used to generate patients' average rating on performing activities of function that are important to them and which were impacted by their neurological condition (Stratford et al. 1995, Mathis et al. 2019, Evensen et al. 2020)
- Mean differences were provided for comparison with minimal clinically important differences (MCIDs), defined as EQ-Index≥0.10, EQ-VAS≥10, and PSFS≥2.8 points (Chen et al. 2016, Mathis et al. 2019)

**Supplementary Table 1.9: NROL Patient outcomes**

| | **Evaluation period<br>Regional<br>Apr 22 – Mar 2024** | | | | | | |
|---|---|---|---|---|---|---|---|
| **Measure** | **N** | **Median entry (IQR)** | **Median exit (IQR)** | **p-value[a]** | **Mean difference (SD)** | **% NROL participants who improved or remained stable (change score (exit-entry) ≥ 0)** | **% NROL participants met or exceeded MCID** |
| EQ-Index | 280 | 0.537 (0.315-0.689) | 0.605 (0.434-0.728) | **<0.001** | 0.070 (0.203) | 64% | 39% |
| EQ-VAS | 280 | 60 (50-74) | 60 (50-75) | **0.009** | 3.03 (19.02) | 68% | 36% |
| PSFS | 282 | 3.33 (2.33-4.67) | 5.00 (3.67-6.75) | **<0.001** | 1.62 (2.03) | 80% | 30% |

[a]Wilcoxon-Signed Rank Test. MCID = Meaningful clinically important difference
