## Supplemental Tables 2 for "Can we enhance neurorehabilitation through regional implementation of group-based telerehabilitation? A mixed methods evaluation of NeuroRehabilitation OnLine (NROL)"

**Supplementary file 2: NROL staff and patient surveys**

Includes:
- NROL staff survey
- NROL patient survey
- Supplementary Table 2.1: Staff and patient respondent characteristics
- Supplementary Figure 2.1: Visual summary of quantitative survey responses (excl. characteristics)

**NROL staff survey**

1. Please indicate your experience of NROL to date (tick all relevant):
   - No experience
   - Have referred patients to NROL
   - Have observed NROL sessions
   - Have helped deliver NROL sessions
   - Have helped coordinate NROL

2. How would you rate your level of proficiency with digital technology before NROL observation or involvement?
   - Non-existent
   - Novice
   - Moderate
   - Expert

3. How would you rate your level of proficiency with digital technology after NROL observation or involvement?
   - Non-existent
   - Novice
   - Moderate
   - Expert

4. Please indicate how much you agree or disagree with the following statements:

| | Strongly disagree | Disagree | No opinion | Agree | Strongly agree |
|---|---|---|---|---|---|
| I feel positive about NROL | ○ | ○ | ○ | ○ | ○ |
| It is clear to me *how* NROL helps to improve rehabilitation delivery for patients with brain injury or stroke | ○ | ○ | ○ | ○ | ○ |
| Supporting the delivery of NROL in my service is acceptable to me | ○ | ○ | ○ | ○ | ○ |
| I feel confident to support the delivery of NROL in my service | ○ | ○ | ○ | ○ | ○ |
| Offering NROL increased the intensity of rehabilitation for our patients | ○ | ○ | ○ | ○ | ○ |
| NROL did not require a lot of effort to implement in my service | ○ | ○ | ○ | ○ | ○ |
| Supporting the delivery of NROL easily fits in with my other priorities | ○ | ○ | ○ | ○ | ○ |
| The effort to implement NROL was worth the benefits | ○ | ○ | ○ | ○ | ○ |
| NROL improves outcomes for patients with brain injury or stroke | ○ | ○ | ○ | ○ | ○ |
| Offering NROL reduced the travel requirements for my team | ○ | ○ | ○ | ○ | ○ |
| I think NROL is a fair and equitable way to provide rehabilitation to patients with brain injury or stroke | ○ | ○ | ○ | ○ | ○ |
| NROL benefitted my professional development | ○ | ○ | ○ | ○ | ○ |
| NROL helped my networking with other staff | ○ | ○ | ○ | ○ | ○ |

5. Please tell us the NHS Trust you work for:
   - East Lancashire Hospitals NHS Trust
   - University Hospitals of Morecambe Bay NHS Foundation Trust
   - Blackpool Teaching Hospitals NHS Foundation Trust
   - Lancashire and South Cumbria NHS Foundation Trust
   - Other

6. Please tell us your work role (tick all relevant):
   - Administrator
   - Assistant
   - Service Manager
   - Occupational Therapist
   - Physiotherapist
   - Psychologist
   - Speech and Language Therapist
   - Technology Support
   - Other (state)

7. What is your job band?
   - 3
   - 4
   - 5
   - 6
   - 7
   - 8

8. What are the benefits of NROL?

9. What are the challenges of NROL?

10. If other regions wanted to do NROL:
    a) What are the key parts of NROL they must include?
    b) What are the key lessons you would pass on?

11. Is there any other feedback about NROL?

**NROL patient survey**

1. Before starting NROL how would you rate your computer and technology skills?
   Non-existent
   Novice
   Moderate
   Expert

2. How would you rate your computer and technology skills after NROL?
   Non-existent
   Novice
   Moderate
   Expert

3. Please indicate how much you agree or disagree with the following statements:

| | Strongly disagree | Disagree | No opinion | Agree | Strongly agree |
|---|---|---|---|---|---|
| Before starting NROL the idea of taking part appealed to me | ○ | ○ | ○ | ○ | ○ |
| I thought NROL was a valuable part of my rehabilitation | ○ | ○ | ○ | ○ | ○ |
| I liked that NROL was an online activity | ○ | ○ | ○ | ○ | ○ |
| I would prefer online rehabilitation rather than face-to-face contact | ○ | ○ | ○ | ○ | ○ |
| NROL was convenient from a time and energy point of view | ○ | ○ | ○ | ○ | ○ |
| I was able to apply what I learnt during NROL | ○ | ○ | ○ | ○ | ○ |
| The peer-support I received from fellow patients was important to me | ○ | ○ | ○ | ○ | ○ |
| The fact NROL was a group activity was important to me | ○ | ○ | ○ | ○ | ○ |
| Working in a group helped increase my confidence | ○ | ○ | ○ | ○ | ○ |
| I was able to learn from the experiences of others | ○ | ○ | ○ | ○ | ○ |
| I had all the equipment and space I needed to allow me to take part in NROL | ○ | ○ | ○ | ○ | ○ |
| I received all the support I needed to access NROL online | ○ | ○ | ○ | ○ | ○ |
| The staff that ran NROL sessions paid attention to my needs | ○ | ○ | ○ | ○ | ○ |

4. Can you give one thing you liked about NROL?

5. Can you give one thing we can do to improve NROL?

6. Please can you tell us what you thought about being in groups with other people?

7. Any other comments.

8. Would you recommend NROL to another person? Yes / No

.

**Table: Staff and patient survey respondent characteristics**

| | **Characteristic** | | % responses |
|---|---|---|---|
| **Staff (n = 49)** | Organisation | ELHT | 47% |
| | | UHMB | 22% |
| | | BTH | 10% |
| | | LSCFT | 18% |
| | | Unspecified | 2% |
| | Role | Occupational Therapist | 29% |
| | *(all relevant)* | Physiotherapist | 24% |
| | | Psychologist | 8% |
| | | Speech and Language Therapist | 10% |
| | | Therapy Assistant/Trainee | 14% |
| | | NROL Operational team | 6% |
| | | Service Manager | 4% |
| | | Unspecified | 6% |
| | Seniority | Band 3 | 6% |
| | | Band 4 | 12% |
| | | Band 5 | 10% |
| | | Band 6 | 45% |
| | | Band 7 | 12% |
| | | Band 8 | 12% |
| | | Unspecified | 2% |
| | Experience with NROL | No direct | 12% |
| | *(all relevant)* | Referred | 57% |
| | | Observed | 57% |
| | | Delivered | 55% |
| | | Coordinate | 24% |
| **Patients (n = 176)** | Organisation | ELHT | 48% |
| | | UHMB | 26% |
| | | BTH | 11% |
| | | LSCFT | 10% |
| | | Unspecified | 6% |
| | Sex | Male | 53% |

**Figure: Visual summary of quantitative survey results**

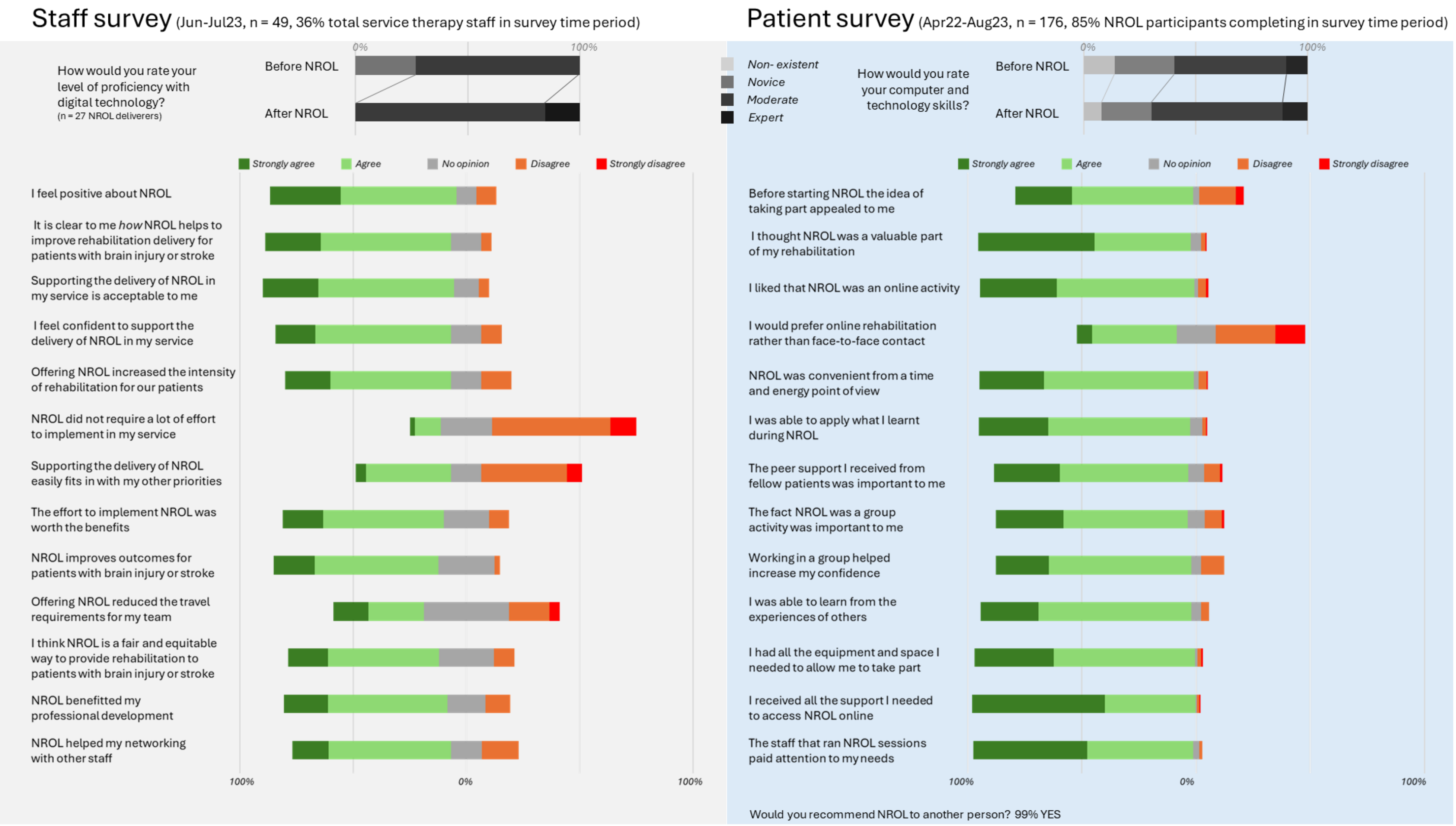
